# Deficient auditory gamma-BOLD coupling in schizophrenia is related to sensory gating deficits

**DOI:** 10.1101/2021.08.31.21262929

**Authors:** Michael S. Jacob, Kaia Sargent, Brian J. Roach, Elhum A. Shamshiri, Daniel H. Mathalon, Judith M. Ford

## Abstract

**Background:** Schizophrenia is associated with aberrant gamma band power, hypothesized to reflect imbalance in the excitation-inhibition (E/I) ratio and undermine neural signal efficiency. Relationships between resting-state gamma, E/I balance, and regional hemodynamics from the fMRI BOLD signal are unknown.

**Methods:** We recorded simultaneous EEG-fMRI at rest, with eyes open, in people with schizophrenia (n= 57) and people without a psychiatric diagnosis (n= 46) and identified gamma and aperiodic EEG parameters associated with E/I balance. Measures from all EEG channels were entered into a whole-brain, parametric modulation analysis followed by statistical correction for multiple comparisons. Sensory gating was assessed using the Sensory Gating Inventory, and psychotic symptoms were assessed using the Positive and Negative Syndrome Scale.

**Results:** Across groups, gamma power modestly predicts a steeper aperiodic slope (greater inhibition), without group differences in either gamma power or aperiodic slope. In schizophrenia, gamma-BOLD coupling was reduced in bilateral auditory regions of the superior temporal gyri and inversely correlated with sensory gating deficits and symptom severity. Analysis of the spectral features of scanner sounds revealed distinct peaks in the gamma range, reflecting a rapidly repeating scanner pulse sound present throughout the resting state recording.

**Conclusion:** Regional hemodynamic support for putative inhibitory and excitatory contributions to resting EEG are aberrant in SZ. Deficient gamma coupling to auditory BOLD may reflect impaired gating of fMRI-scanner sound.

## Introduction

Signal processing aberrancy in schizophrenia (SZ) spans sensory (Javitt and Sweet, 2015), motor (Morrens et al., 2014) and cognitive domains (Guo et al., 2019). Rather than reflecting a singular neuroanatomical deficit or specific network dysfunction, these composite abnormalities point toward global brain dysfunction (Yang et al., 2014), potentially arising from a fundamental excitatory-inhibitory (E/I) imbalance (Sohal and Rubenstein, 2019). Post-mortem studies in SZ and cellular studies in animal models have identified E/I imbalance owing to impairments in GABA and glutamate synthesis (McGeer, 1977), receptor expression and function (Benes et al., 1992; Olney and Farber, 1995), and morphological features of interneurons (Blum and Mann, 2002; Kaar et al., 2019; Sullivan et al., 2018). Relating cellular abnormalities to network and human level neurophysiology in patients with SZ remains a central translational challenge, necessary to define etiologic mechanisms, develop novel diagnostic markers and guide novel treatments. A necessary first step is to test human biomarkers that have been linked to underlying neurophysiology.

Among potential biomarkers recorded from electroencephalography (EEG), periodic activity in the gamma range and broadband, aperiodic activity have both shown promise for bridging human signaling deficits with cellular measures of neural activity. Abnormal gamma rhythms are a prominent features of SZ (Roach and Mathalon, 2008; Shin et al., 2011), with reductions in amplitude and phase-locking reported in both evoked and resting-state conditions (for review see: Uhlhaas and Singer, (2010), but see White and Siegel (2016) for evidence of *increased* gamma power during rest). Gamma rhythms are generated through coordinated interactions between excitatory and inhibitory currents (Atallah and Scanziani, 2009; Buzsáki and Wang, 2012), principally mediated by inhibitory interneurons (Sohal and Rubenstein, 2019) that may be impaired in SZ (Lewis et al., 2012). Alterations in gamma power in SZ are therefore thought to relate to disturbances in inhibitory transmission and E/I balance (Chung et al., 2016; Grent-’t-Jong et al., 2018; Selten et al., 2018; Sohal et al., 2009). In ERP and EEG studies of SZ patients, gamma abnormalities are linked to stimulus specific processing deficits, as seen in auditory paradigms (Kwon et al., 1999; Roach and Mathalon, 2008; Tada et al., 2020) that are linked to impaired sensory gating (Nguyen et al., 2020) and auditory hallucinations (Kuga et al., 2016). The corresponding neuroanatomy of resting-state gamma power in SZ is largely unknown.

Recent work has demonstrated that E/I balance may also be inferred from the slope of the local field potential (LFP) or EEG power spectrum (Gao et al., 2017; Podvalny et al., 2015; Poil et al., 2012). The spectral slope reflects aperiodic (non-oscillatory) EEG activity that follows a power law (1/f) distribution, comprising the largest proportion of EEG power (He et al., 2014). Gao et al., (2017) show that increasing synaptic inhibition, through computational models, and experimentally through the administration of a GABAergic anesthetic, results in a significant increase (steepening) of the aperiodic slope. In contrast with gamma rhythms, which tend to reflect local processing dynamics, human studies of aperiodic slope tend to capture global brain states such as the level of consciousness (Colombo et al., 2019) or sleep (Ma et al., 2018), with greater alertness associated with a flatter slope. This is consistent with emerging cellular research that E/I balance fluctuates over time and is related to arousal (Bridi et al., 2020; Lendner et al., 2020).

As a global measure of excitability, the aperiodic slope may index criticality dynamics that enable rapid state-shifts across regions, including the emergence of resting-state networks (Haimovici et al., 2013). Therefore, E/I imbalance in SZ may impair coordination across or between brain networks as revealed by well studied BOLD resting-state abnormalities (Allen et al., 2019; Manoliu et al., 2014; Menon, 2011; Palaniyappan et al., 2013; Pettersson-Yeo et al., 2011; Whitfield-Gabrieli and Ford, 2012). While preliminary evidence suggests aperiodic changes are present in SZ (Peterson et al., 2018), but see (Racz et al., 2021), the neuroanatomy associated with aperiodic EEG in SZ remains unknown. Our goals are twofold: first, to test for group differences in aperiodic spectral slope in SZ and second, to examine the relationship between resting-state EEG markers of E/I balance with regional BOLD activity in SZ.

Even at “rest,” neural activity may be driven by background stimuli that are unavoidable, such as scanner background sound (Benjamin et al., 2010) and that may also drive arousal (Kobald et al., 2016). We have previously hypothesized that EEG measures of E/I balance might reflect processing of scanner background sound and spontaneous fluctuations in arousal during the resting-state (Jacob et al., 2021). In support of this hypothesis we have observed that in healthy participants, gamma power fluctuations were associated with BOLD activity in locally circumscribed auditory regions, whereas aperiodic power fluctuations were associated with distributed networks, including auditory, cerebellar, salience and prefrontal regions (Jacob et al., 2021).

In this current study, we hypothesized that E/I imbalance in SZ might be revealed from reduced gamma power (deficient local inhibitory current) and flatter aperiodic slope (globally deficient inhibition) and associated with atypical regional and distributed BOLD signals. Given our previous finding of aperiodic and gamma EEG linked to auditory-salience regions, we further hypothesized that impaired gamma and aperiodic activity in SZ would be associated with subjective impairment in the gating of environmental auditory stimuli. Auditory gating deficits have been well-studied in schizophrenia (Hamilton et al., 2018; Hetrick et al., 2012; Mayer et al., 2013; Nguyen et al., 2020; Tregellas et al., 2009) and might uniquely contribute to and/or alter resting-state neural activity in the context of MRI scanner sound.

Using simultaneous EEG-fMRI, we first measure aperiodic and gamma EEG signals in healthy participants and SZ participants while at rest and then use those signals as predictors in a whole-brain, parametric modulation analysis to test for group differences in EEG-BOLD coupling. Contrary to our initial hypothesis, we find no group differences in resting-state gamma or aperiodic EEG. However, we do find reduced BOLD coupling to gamma EEG in regions of the auditory cortex in SZ that are associated with impaired sensory gating. Our results have implications for fMRI resting-state paradigms and studies of SZ in particular: changes in “resting-state” activity may be driven by unavoidable scanner background stimuli. Rather than exclusively viewing scanner background sound as a confounding variable, we suggest that the MR-scanner environment might be considered a semi-naturalistic paradigm to study “background” fluctuations in neural excitability that might contribute to sensory processing impairments in schizophrenia.

## Methods

### Participants

The study included 57 people with schizophrenia (n = 45) and schizoaffective disorder (n = 12, collectively referred to as SZ), and 46 healthy participants (HP) matched for age and gender. One SZ participant and two HP participants had no usable EEG data collected during fMRI recording due to an amplifier connection problem and were excluded from the analysis. Participants were recruited through advertising and word of mouth, and SZ participants were referred by community outpatient clinics. Clinical and demographic data are presented in Table 1. The healthy participant sample and analysis has been previously reported (Jacob et al., 2021).

**Table 1.**
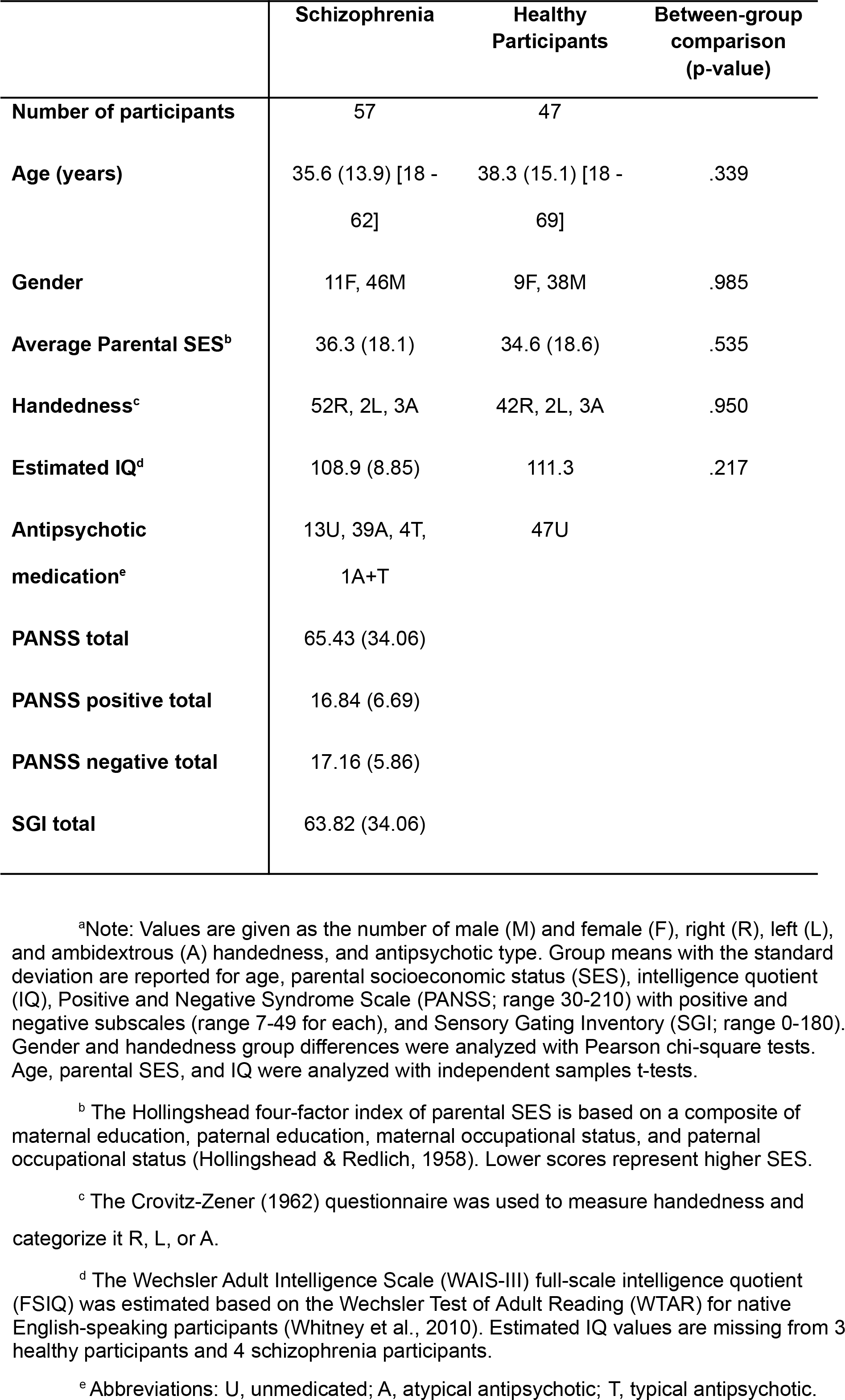
Group Demographic Data^a^

For both groups, exclusion criteria included substance abuse in the past 3 months, any significant medical or neurological illness, or head injury resulting in loss of consciousness. In addition, HP were excluded for any current or past psychiatric disorder based on the SCID-IV, any history of substance dependence (except nicotine), or having a first-degree relative with a psychotic disorder. SZ participants were excluded if they met criteria for substance dependence within the last year. A trained research assistant, psychiatrist, or clinical psychologist conducted all diagnostic and clinical interviews, including the SCID-IV (First et al., 1995) and the Positive and Negative Syndrome Scale (Kay et al., 1987). The Sensory Gating Inventory (Hetrick et al., 2010) was completed as a written questionnaire.

Study procedures were approved by the University of California at San Francisco and the San Francisco Veterans Affairs Medical Center Institutional Review Boards. All participants provided written informed consent.

### EEG-fMRI acquisition

Simultaneous EEG-fMRI data were acquired during rest. Participants were instructed to keep their eyes open and fixate on a white cross displayed on a black screen for 6 minutes. Acquisition methods are discussed in detail elsewhere (Ford et al., 2016; Jacob et al., 2021) and discussed in brief below. Structural and functional MRI data were collected using a 3T Siemens Skyra scanner. The structural imaging protocol was a magnetization-prepared rapid gradient-echo (MPRAGE) T1-weighted high-resolution image (2300 ms TR, 2.98 ms TE, 1.20 mm slice thickness, 256 mm field of view, 1.0 × 1.0 × 1.2 voxel size, flip angle 9°, sagittal orientation, 9:14 min). The fMRI protocol was an AC-PC aligned echo planar imaging (EPI) sequence (2000 ms TR, 30 ms TE, flip angle 77°, 30 slices collected sequentially in ascending order, 3.4 × 3.4 × 4.0 mm voxel size, slice gap=1mm, Field of View=220 mm, 182 frames, 6:08 min).

### fMRI Preprocessing

#### Statistical Parametric Mapping 8

(SPM8; http://www.fil.ion.ucl.ac.uk/spm/software/spm8/) was used for image preprocessing. Motion correction was performed via affine registration where each image was realigned to the first image of the run, and images were slice-time corrected with respect to the middle slice to adjust for timing differences within each TR. We then implemented aCompCor (anatomic component based noise correction), a principal components-based approach for noise reduction of fMRI data by identifying “noise” components from weighted averages of white matter and CSF voxel time series. (Behzadi et al., 2007).

### EEG Acquisition

EEG data were collected from 32 scalp sites with an additional electrode placed on the lower back to monitor ECG. We used sintered Ag/AgCl ring electrodes in an MR-compatible electrode cap from Brain Products (Munich, Germany) according to the 10-20 system of electrode placement. Electrode impedances were kept below 10 kΩ. FCz was used as reference and AFz as ground. The data were recorded with a bandpass filter of 0.01–250 Hz and digitized at a rate of 5 kHz with 0.5 μV resolution (16 bit dynamic range, 16.38 mV). The nonmagnetic EEG amplifier was placed in the scanner bore behind the head coil, and the participant’s head was immobilized using cushions. EEG data were transmitted to a BrainAmp USB adapter and synchronized to the MRI master clock via a SyncBox (Brain Products).

### EEG Preprocessing

MR gradient artifacts were removed using artifact subtraction proposed by a (Allen et al., 2000) and implemented in Brain Vision Analyzer 2.0.4.368 software (BrainProducts). The correction involved subtracting an artifact template from the raw data, using a baseline-corrected sliding average of 21 consecutive volumes to generate the template (Allen et al., 2000). EEG data were then down-sampled to 250 Hz.

ECG data were used to determine when heartbeats occurred in the EEG signal. Semi-automatic heartbeat detection was performed in Brain Vision Analyzer 2 (BrainProducts, Gilching, Germany), where heartbeats were identified in the ECG channel that exhibited a high temporal correlation (r > 0.5) with a heartbeat template (a heartbeat chosen by the algorithm occurring about 30 s into the run) and within-range amplitude (0.6– 1.7 μV). Trained research assistants adjusted templates and search windows, manually identifying pulses as needed. Ballistocardiac artifact removal was achieved in the same manner as gradient artifact removal (Allen et al., 1998; Schulz et al., 2015; Shams et al., 2015).

We implemented canonical correlation analysis (CCA) as a blind source separation technique to remove electromyographic noise, using a method identical to previous reports (Baenninger et al., 2017; Mathalon et al., 2018) and similar to that used by others (De Clercq et al., 2006; Ries et al., 2013).

Single trial data were re-referenced to an average reference and data were obtained from the prior reference channel, FCz. Artifactual TR intervals were rejected with FASTER criteria (Nolan et al., 2010). Independent components analysis (ICA) was then performed on each participant in EEGLAB (Delorme & Makeig, 2004), generating 32 independent components. Noise components were identified by FASTER criteria as well as spatial correlations (r >0.8) with eyeblink and ballistocardiac artifact templates. The eyeblink template was created by averaging components resembling frontal-only activity, while the ballistocardiac template was created by setting midline electrodes to 0 and the two hemispheres with opposing signs, resembling a dipole effect. These noise components were then removed from the data during back-projection. Finally, outlier channels within epochs were interpolated with spherical spline interpolation.

### EEG Parameter Estimation

Data from all 32 channels were segmented into 2-second epochs corresponding to whole brain acquisition time for each volume. We computed a Fourier transform within each epoch with a 2-second Hanning window (frequency resolution of 0.5 Hz) to obtain a power spectrum for each epoch. For each resulting power spectrum, we isolated periodic and aperiodic components using FOOOF within the range of 1-50 Hz. (Fitting Oscillations and One Over F, https://fooof-tools.github.io/fooof/, (Donoghue et al., 2020). Briefly, FOOOF estimates a first-pass least-squares fit to obtain initial seed values for the aperiodic spectral offset and spectral exponent. This fit is subtracted from the original power spectrum, creating a flattened spectrum. The lowest amplitude points (below the 2.5th percentile) are then used in a final estimate of the exponent and offset values.

The final aperiodic fit is then subtracted from the power spectrum to isolate residual periodic power. We examined low gamma oscillations within a band range of 30-50 Hz that are consistent with definitions provided by the International Federation of Clinical Neurophysiology (Babiloni et al., 2020) and prior simultaneous EEG-fMRI studies (Mantini et al., 2007). Mean power values for gamma and the aperiodic spectral exponent were extracted and entered into a parametric modulation analysis, discussed below.

### Parametric Modulation Analysis

The details of our approach are reported in a prior manuscript (Jacob et al., 2021). Time series for gamma power and the aperiodic exponent were obtained for each 2 sec TR, and then z-scored. We employed a whole-brain, parametric modulation analysis (Büchel et al., 1998) using fluctuations in gamma and aperiodic EEG time series as predictors for BOLD dynamics (Laufs et al., 2003) in first-level fMRI analyses in SPM8. aCompCor noise components and motion parameters were included as nuisance regressors. For each gamma and aperiodic EEG parameters, beta coefficients were estimated for each voxel’s time series, resulting in beta images representing BOLD fluctuations predicted by EEG parameter variance. Beta images were normalized to the Montreal Neurological Institute’s EPI template (http://www.bic.mni.mcgill.ca) and smoothed with a 6 mm Gaussian kernel.

Second-level group models identified EEG-fMRI clusters that showed significant differences in gamma and aperiodic BOLD coupling between SZ and HP groups. Our initial voxelwise cluster-finding threshold was set to p=0.001 (two-tailed) and a spatial extent of 10 voxels. For all clusters surviving the initial threshold across any of the 32 channels, cluster-level p-values were FDR corrected to account for multiple comparisons. For remaining clusters, beta coefficients were extracted and correlated with clinical measures to assess the relationship between EEG-BOLD coupling and symptoms. Significant clusters were identified from electrodes FC5 and T8. Electrode FC5 was selected for EEG-only analysis of gamma and aperiodic slope; the results do not change for T8 or other electrodes.

## Results

### Gamma and Aperiodic Slope

There were no group differences in gamma power or aperiodic slope in EEG alone (gamma: t(101) = .002, p = .999; aperiodic slope: t(101) = -.872, p = .385; see Figure 1a). Across all participants, greater gamma power was associated with steeper slope (greater inhibition) over all TR intervals (r=0.44, p<0.00001; see Figure 1b).

**Figure 1.**
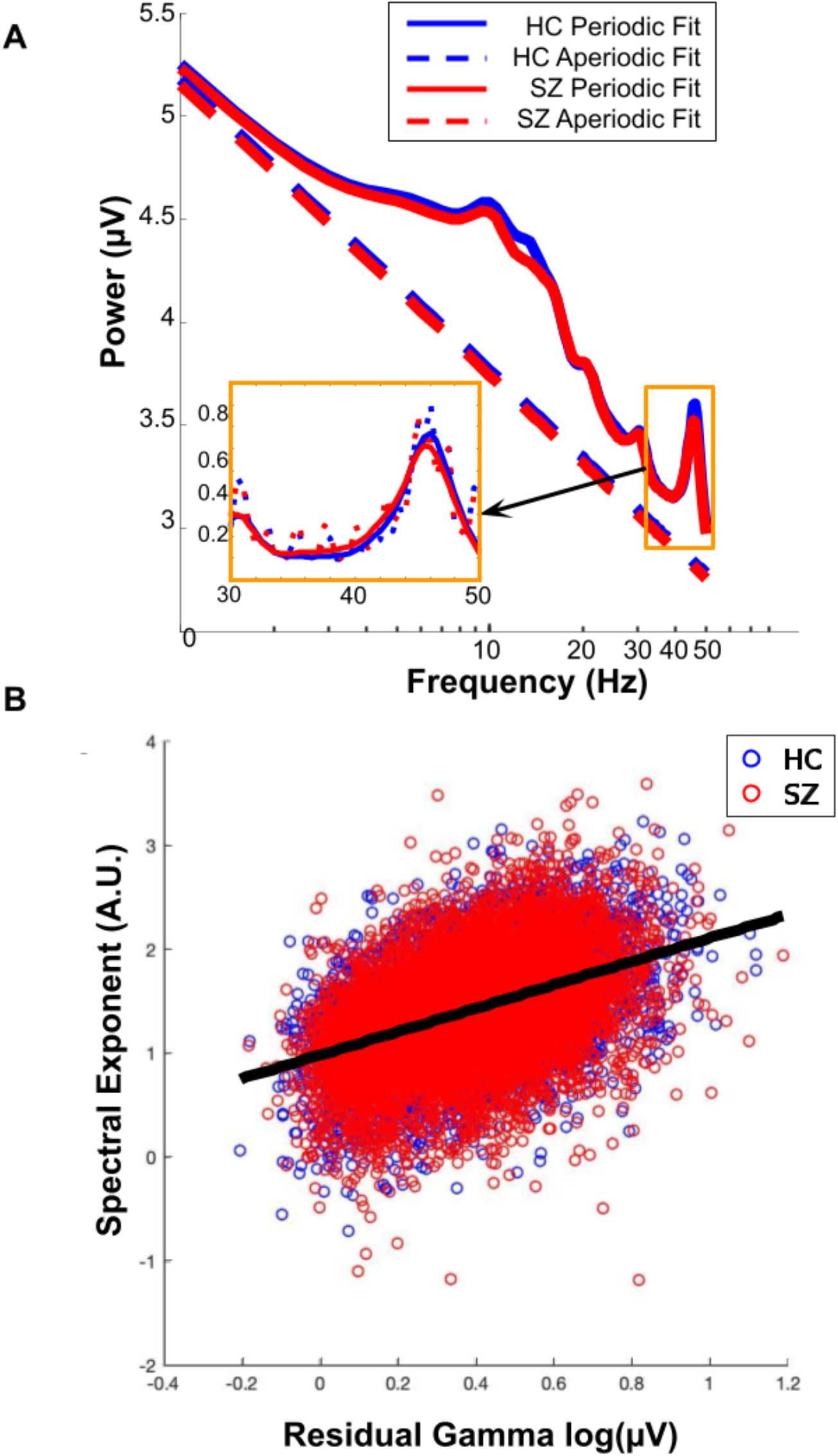
**(A)** Grand average spectra, derived from the FOOOF aperiodic (dashed line) and periodic fit (solid line) for all SZ (red) and healthy participants (blue) from electrode FC5. Orange Inset: Average residual gamma power after subtracting the aperiodic fit from the periodic fit **(B)** Correlation between residual gamma and the aperiodic exponent over all TR intervals and across groups from electrode FC5.

### EEG-BOLD Coupling

Whole-brain parametric modulation analyses revealed 3 clusters with an aberrant pattern of EEG-BOLD coupling in SZ. Gamma-BOLD coupling from two regions of the bilateral superior temporal gyri was reduced in SZ (Figure 2). In addition, BOLD activity in the left superior frontal gyrus (including supplementary motor area) showed an opposite direction of coupling in HP and SZ groups (Figure 3). In this cluster, HPs showed BOLD activity that was negatively coupled to the aperiodic exponent (flatter slope, more BOLD) whereas SZ showed BOLD activity that was positively coupled to the aperiodic exponent (steeper slope, more BOLD). Table 2 shows cluster statistics and MNI coordinates of all significant clusters, FDR corrected for 32 EEG channels.

**Table 2.**
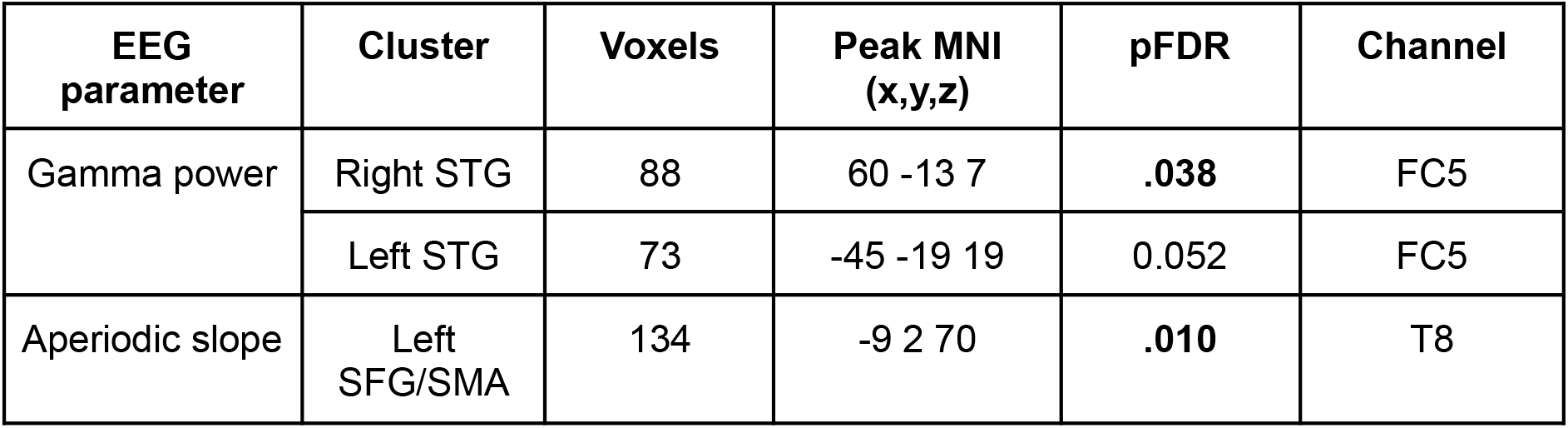
Clusters with a significant difference in EEG-BOLD coupling between SZ and HP

**Figure 2.**
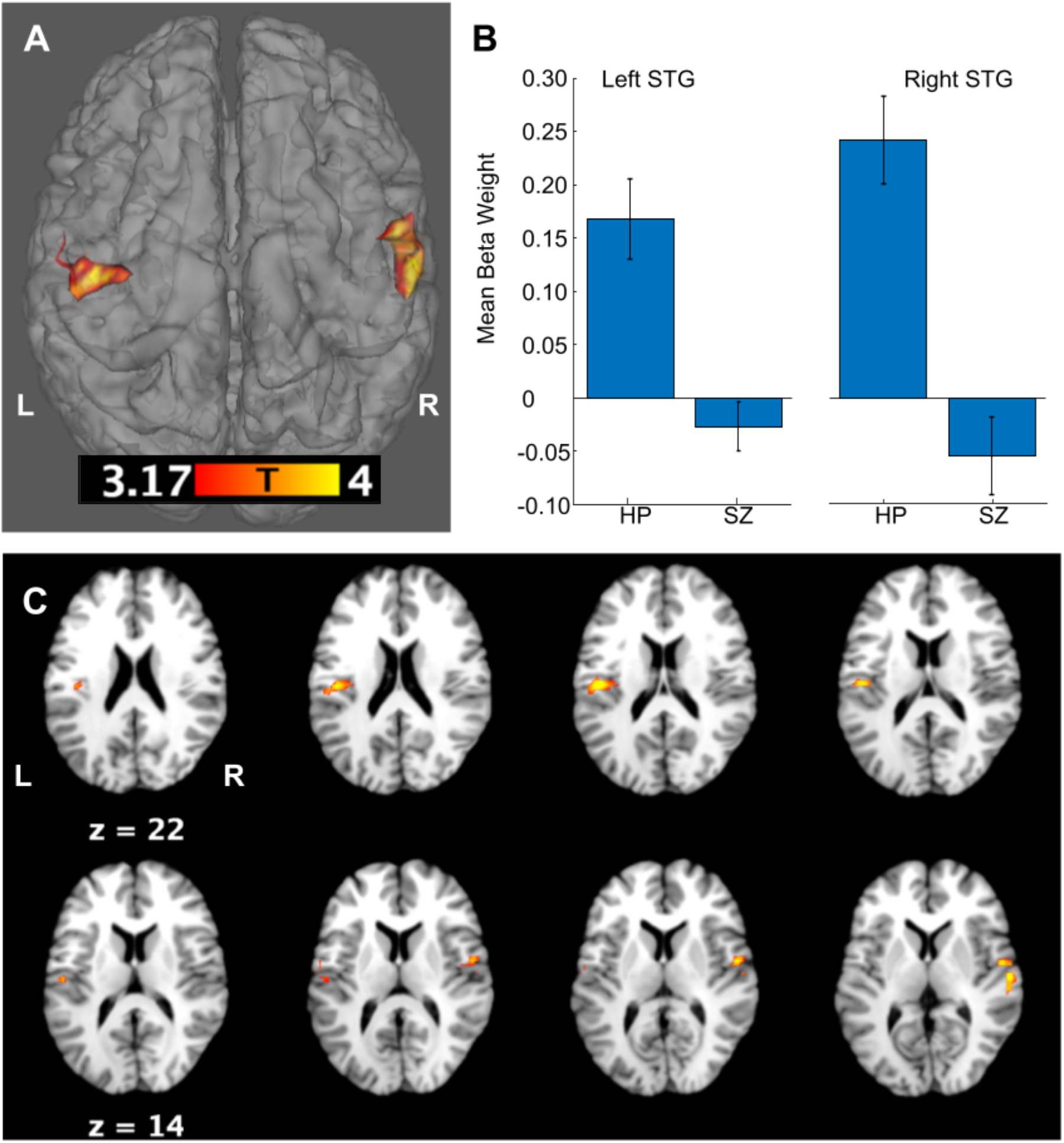
(A) Two clusters identified with deficient BOLD Coupling to Gamma power in SZ (HP > SZ). (B) Beta weights derived from the clusters shown in (A) for the left STG cluster (left) and right STG cluster (right) for healthy (HP) and schizophrenia (SZ) participants. (C) Axial sections of the clusters shown in (A) reveal bilateral coupling in posterior insula and auditory regions of the STG.

**Figure 3.**
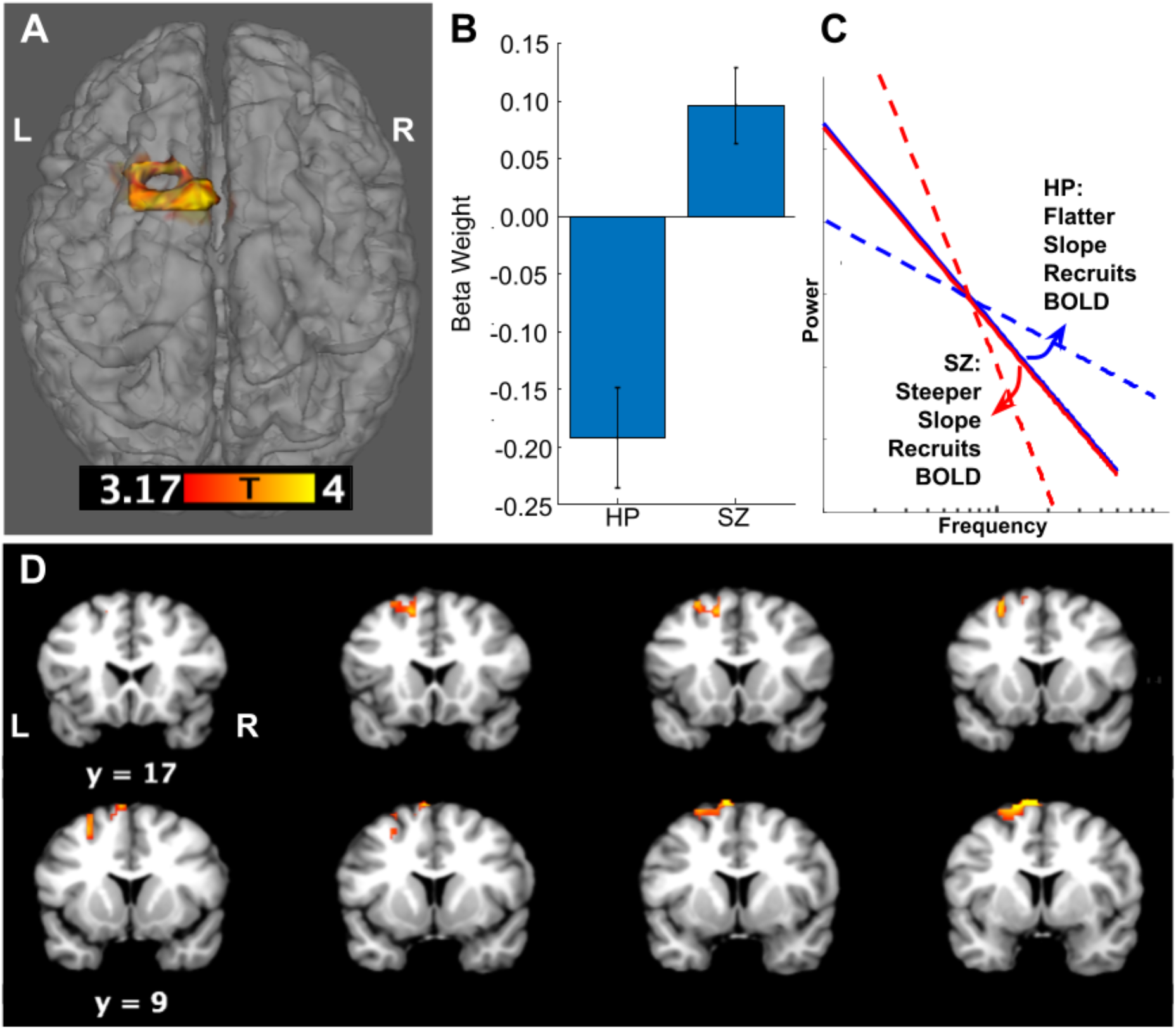
(A) One cluster in the superior frontal gyrus identified an opposite pattern of coupling with the aperiodic exponent in HP and SZ participants (SZ > HP). (B) Beta weights derived from the cluster shown in (C) reveal positive coupling in SZ and negative coupling in HPs. (C) A schematic indicates the direction of coupling effects, with positive coupling indicating a BOLD response associated with a steeper slope (a greater spectral exponent)and negative coupling indicating a BOLD response associated with flatter slope (smaller spectral exponent). (D) Coronal sections of the cluster shown in (A) reveals coupling differences in the supplementary motor area.

**Figure 4.**
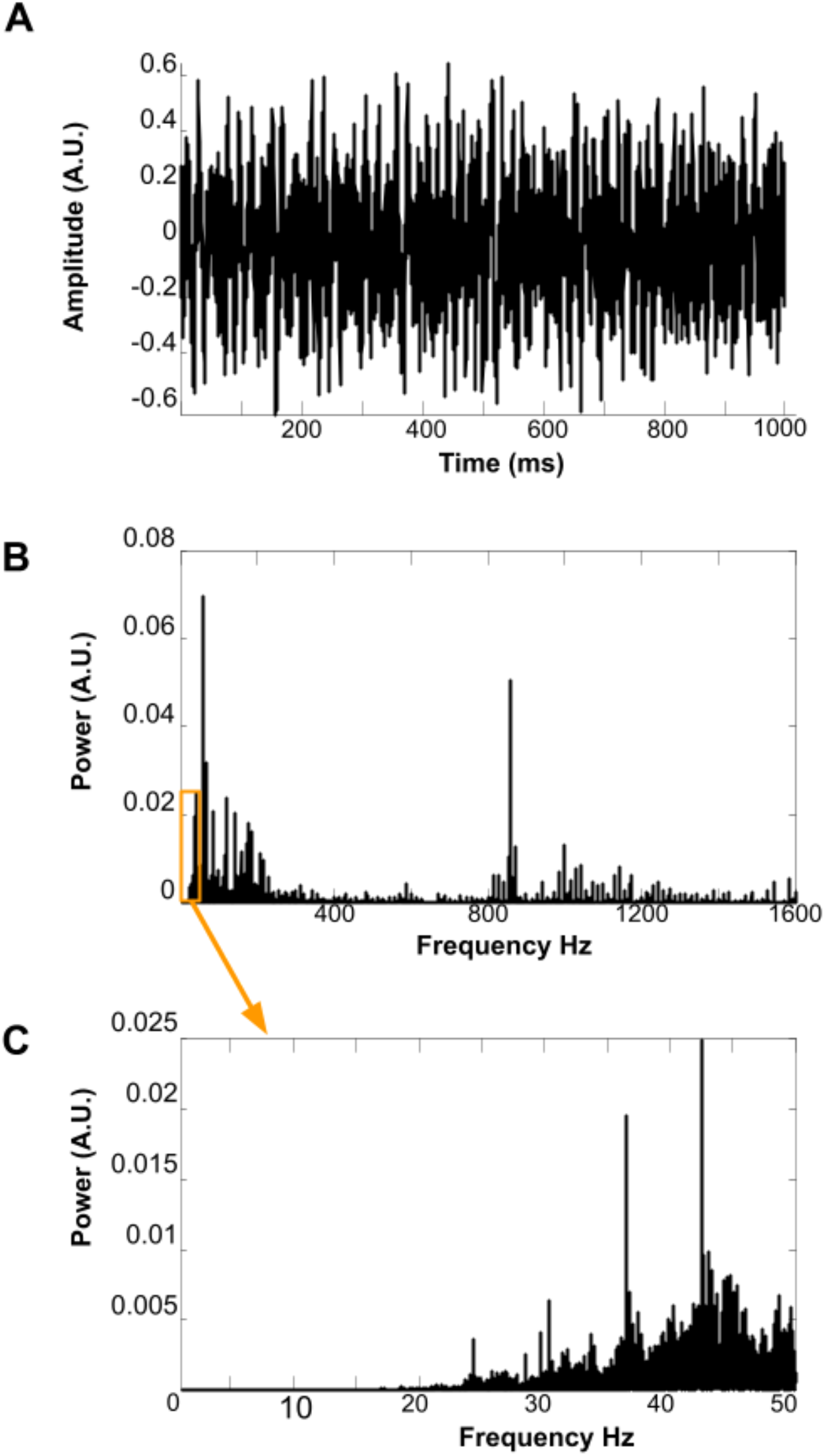
(A) Tracing of the amplitude of the scanner sound signal recorded during the first second of the resting-state sequence. (B) Sound signal spectra of the entire 6 min resting state sequence. (C) Spectra within the range of EEG activity recorded in this experiment.

### Scanner Sound Spectra

Given the presence of strong gamma-BOLD coupling in the auditory cortex with a distinct gamma peak between 40-50 Hz (see Figure 1a), we asked whether gamma-BOLD coupling in this region might reflect auditory processing of scanner background sound. In a preliminary test of this hypothesis, we recorded scanner sound during the resting-state sequence and computed the power spectral density. We observed distinct low frequency peaks between 30-100 Hz in the power spectrum reflecting multiple, rapidly repeating scanner pulse sounds at ∼900 Hz (carrier frequency, see Figure 5B). Several distinct spectral peaks were present in the range of our EEG analysis, particularly at ∼45 Hz in the gamma range (see Figure 5C), reflecting a repeating scanner pulse sound that might trigger EEG entrainment.

**Figure 5.**
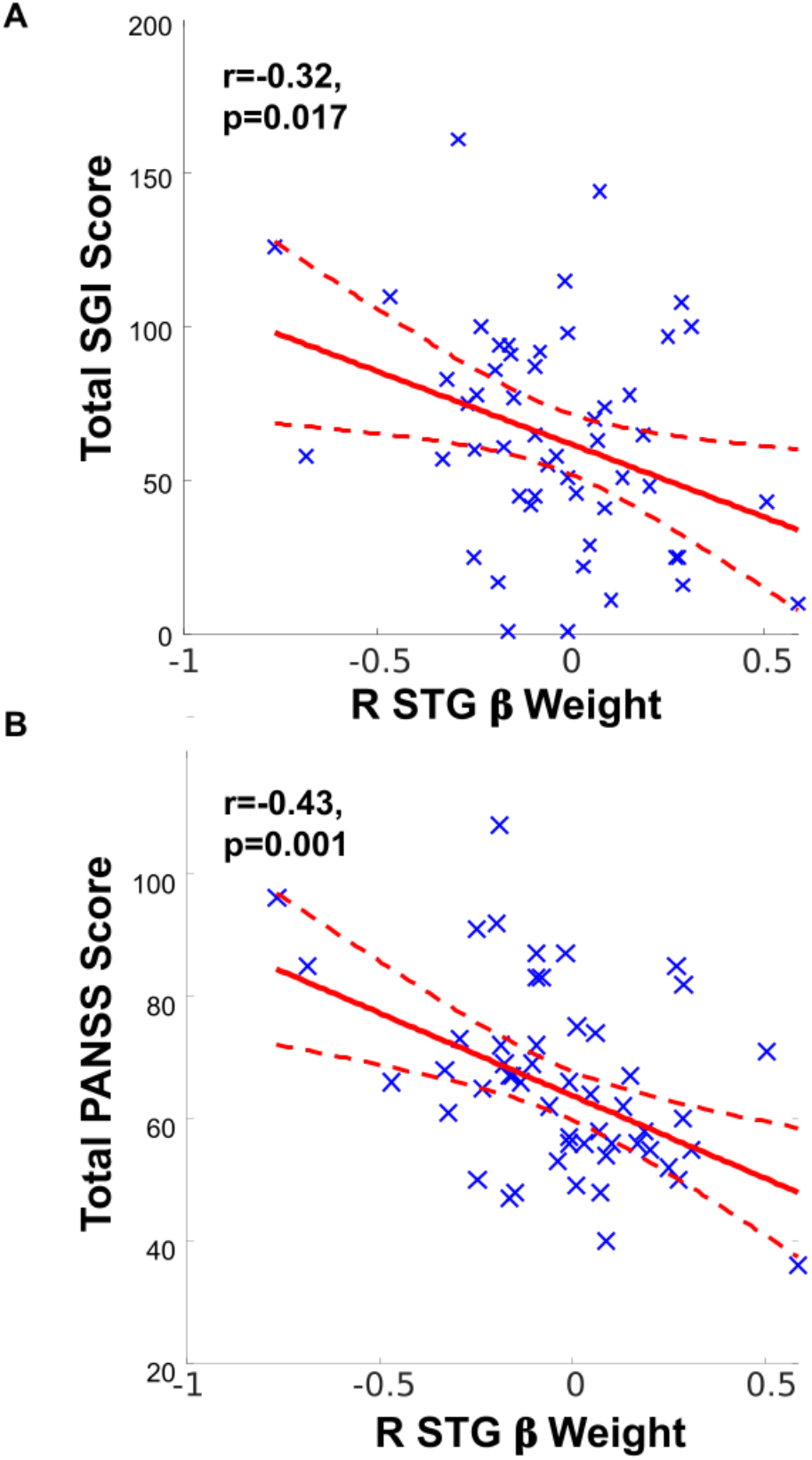
(A) Total sensory gating inventory score inversely correlates with the strength of R STG BOLD-Gamma coupling SZ (B) Greater psychotic symptoms on the PANSS inversely correlates with the strength of R STG BOLD-Gamma coupling SZ.

### Symptoms and Sensory Gating

Within SZ, beta weights extracted from the gamma-BOLD coupling in STG clusters inversely correlate with subjective self-report of sensory gating deficits as measured by the SGI (see Figure 5A and Table 3). That is, deficient gamma-BOLD coupling in these regions was associated with worse sensory gating. Deficient gamma-BOLD in the R STG was also associated with overall symptom severity and positive symptom severity (see Figure 5B and Table 3).

**Table 3.** Correlations between beta weights extracted from significant clusters and symptom severity.

| EEG parameter | Cluster | PANSS total |  | PANSS positive |  | PANSS negative |  | SGI |  |
| --- | --- | --- | --- | --- | --- | --- | --- | --- | --- |
|  |  | r | p | r | p | r | p | r | p |
| Gamma power | R STG | -.427 | **.001** | -.297 | **.025** | -.253 | .059 | -.320 | **.017** |
|  | L STG | -.416 | **.001** | -.280 | **.035** | -.254 | .059 | -.169 | .178 |
| Aperiodic Slope | L SFG | .079 | .563 | .033 | .807 | .142 | .296 | .075 | .554 |

## Discussion

We investigated the hemodynamic correlates of ongoing E/I balance at rest by examining simultaneous BOLD coupling to fluctuations in gamma power and the aperiodic spectral slope in healthy participants and individuals with SZ. We found no group difference in mean gamma power or aperiodic slope from EEG measures alone. However, we found reduced BOLD coupling to gamma power in superior temporal gyri in individuals with SZ, including regions of the auditory cortex. Patients with worse subjective gating of sensory stimuli and more severe psychotic symptoms showed the most deficient BOLD coupling. SZ participants also showed inverse coupling to aperiodic slope in superior frontal regions, specifically the supplementary motor cortex. We hypothesized that gamma oscillations in our paradigm might be entrained to the sounds generated by the MR scanner. In preliminary support of this hypothesis, we identified peaks in the gamma range from a spectral analysis of the sounds generated during the resting state sequence.

In summary, and across all participants, we find that gamma power and aperiodic slope, two markers of inhibitory conductance, are robustly coupled with BOLD signals in regions associated with auditory processing (see also Jacob et al., (2021)). These findings suggest that while at rest in an MR-scanner, the brain devotes neural activity and recruits considerable metabolic energy to the processing of scanner sound. This finding also suggests that fMRI resting-state paradigms may be an unavoidable but also semi-naturalistic paradigm to study background auditory sound processing in SZ, a population for whom sensory processing deficits are well-known and related to functional impairment (for review see Javitt and Sweet, (2015)). Our findings have implications for studies of resting-state E/I balance, sensory gating and neurometabolic coupling in SZ. In the discussion that follows, we consider these implications in the context of prior studies, including limitations and future directions for this line of research.

### E/I Balance, Gamma and Aperiodic Slope

A priori, we planned to examine gamma oscillations and aperiodic slope as putative indices reflecting E/I balance. Across all participants, gamma power was correlated with aperiodic slope (increased power associated with a steeper, increased slope), explaining about ∼22% of the variance in aperiodic slope. We suggest that gamma power and spectral slope reflect E/I balance across local and global networks, respectively. This perspective is supported by our prior finding of a distributed network of brain regions associated with the spectral exponent (bilateral superior temporal regions, mid-anterior cingulate, cerebellum and regions of the superior frontal gyrus), whereas BOLD associated with gamma power fluctuations are localized to the bilateral superior temporal gyri. A local/global perspective is also consistent with MEG findings that aperiodic activity is widely distributed (Donoghue et al., 2020), while gamma oscillations tend to be narrowly localized according to the general principle that the spatial extent of oscillatory activity is inversely proportional to its frequency (Buzsaki, 2006; Donner and Siegel, 2011; von Stein and Sarnthein, 2000).

In contrast with our hypothesis, we observed no difference in either spectral slope or gamma power between HP and SZ. Atypical gamma oscillations are widely reported in SZ, with most studies reporting reduced power and phase locking of both evoked and resting-state oscillations (see Uhlhaas & Singer, 2010 for review). However, results of resting-state studies are less consistent, with some studies also reporting *increased* gamma power during rest (Grent-’t-Jong et al., 2018); (see White and Siegel (2016) for review). In contrast with gamma band studies, there is limited prior work examining the aperiodic slope in SZ with a preliminary report of reduced spectral exponent during a working memory task (Peterson et al., 2018) and no apparent difference in another study (Racz et al., 2021). Given that we are unable to identify group differences in either of our putative measures of E/I balance in the context of a simultaneous EEG-fMRI study, we suggest that features of the scanner environment, and scanner sound in particular (as we discuss below), may be driving neural excitability. This driving of EEG excitability by background stimuli might obscure spontaneously driven neural activity, which is commonly studied in *in-vitro* models of E/I balance. For awake, human studies of E/I balance, “background” fluctuations in neural excitability might be inherently linked to the environmental context (both internal and external), making it difficult (if not impossible) to truly separate endogenous, spontaneously driven neural activity from extrinsically driven neural stimuli.

### Deficient Gamma-BOLD Coupling

HPs show coupling between gamma power and BOLD activity in the right and left STG, while individuals with SZ show deficient gamma-BOLD coupling in these same regions. These results align closely with our prior examination of EEG-BOLD coupling in SZ using joint independent component analysis in an auditory processing task (Ford et al., 2016). In that study we identified fMRI activation in superior and middle temporal gyri associated with the N100 ERP, a component elicited 100 ms following an auditory stimulus. We find similar activation in superior temporal gyrus associated with gamma power; we therefore suggest that even in the absence of an explicit auditory task, the loud scanner noise may induce STG gamma-BOLD coupling that reflects auditory processing.

Our investigation into the spectral features of the scanner environment revealed a distinct ∼45 Hz peak in the power spectrum reflecting a rapidly repeating scanner pulse sound at ∼800 Hz (carrier frequency). This additional evidence supports our hypothesis that gamma-BOLD coupling in the auditory cortex reflects processing of scanner sound. Reduced STG gamma-BOLD coupling was associated with more severe sensory gating deficits; we speculate that gamma-BOLD coupling in the auditory cortex may reflect the neurophysiological processes involved with suppressing or gating irrelevant scanner sound.

Deficits in basic auditory processing have been extensively reported in SZ (see (Javitt and Sweet, 2015) for review). Gamma power (Spencer et al., 2008) and auditory BOLD activation (Kiehl and Liddle, 2001) are impaired during auditory paradigms in SZ. Most relevant to the current study are auditory steady state response (ASSR) paradigms, in which individuals with SZ show reduced power and phase-locking of gamma oscillations to rapidly repeating (40 Hz) auditory stimuli (see Uhlhaas & Singer, 2010 and Thuné et al., 2016 for review). We speculate that scanner sound during our “resting-state” paradigm may trigger gamma entrainment in a roughly analogous manner to ASSR paradigms. However, it is notable that we did not find reductions in gamma power in SZ during our paradigm. Since scanner sound is not task-relevant, given our resting-state instructions to maintain visual fixation, attentional effects might reduce entrained auditory power (Manting et al., 2020) which could be further impaired in SZ (Javitt and Sweet, 2015). Future studies might employ interleaved “silent” fMRI (Rondinoni et al., 2013) or EEG recordings of fMRI scanner stimuli (Pripfl et al., 2006) to disentangle relationships between evoked gamma, scanner sound and attentional orientation.

### Aperiodic-BOLD Coupling

HP and SZ participants showed opposite BOLD coupling relationships to the aperiodic slope in frontal regions. That is, a larger BOLD signal was associated with a flatter (more excitatory) slope in HPs and a steeper (more inhibitory) slope in SZ. This finding was localized to the SFG and regions of the supplementary motor area (SMA). Notably, these findings overlap spatially with regions associated with auditory ERP components reported in Ford et al., (2016). In that study we found BOLD activation in both auditory cortex and frontal regions including the SFG that was associated with the P200 ERP, which is thought to index later auditory processing modulated by attention. Our finding of altered aperiodic-BOLD coupling in the SFG may relate to attentional mechanisms involved in auditory processing of scanner sound. Individuals with SZ show reduced frontal BOLD recruitment during task-relevant stimuli but allocate excessive resources to task-irrelevant stimuli (Hazlett et al., 2008), that could also be the case with MR scanner sound, and potentially as compensation for deficient sensory gating within more primary auditory regions as we observe here.

The SMA is also involved with auditory processing and particularly auditory imagery (Lima et al., 2016; McGuire et al., 1996); recruited during both self-initiated auditory imagery and spontaneous auditory verbal hallucinations in non-clinical populations (Linden et al., 2011). Participants with SZ have shown reduced grey matter in the SMA that is associated with abnormal motor behavior (Exner et al., 2006; Stegmayer et al., 2014), and our findings suggest an additional interpretation of the SMA linked to auditory/sensory processing.

Broadly, these findings may be interpreted in the context of frontal inhibitory deficits in SZ and compensatory hemodynamic/metabolic support. Converging lines of research indicate that SZ is associated with disturbances in GABA interneurons, particularly in the dorsolateral prefrontal cortex (DLPFC; Blum & Mann, 2002; Lewis et al., 2005). Our findings indicate that frontal metabolic work required to maintain E/I balance functions in *opposite* directions in SZ and HP: increased metabolic work is required for excitation in HP and for inhibition in SZ. Thus, while we did not find evidence of altered aperiodic slope in SZ, we speculate that altered slope-BOLD coupling may be indicative of compensatory metabolic resources required to maintain sufficient frontal inhibition in SZ.

### Neurometabolic Coupling in SZ

That we were able to identify reduced BOLD coupling but unable to identify changes in EEG suggests several possible scenarios related to inhibitory gamma and aperiodic slope: (1) Neural deficit underlies deficient BOLD but these effects were not discernible from surface EEG recordings. (2) A hemodynamic deficit underlies preserved gamma power and slope, or (3) A combination of (1) and (2) that might suggest a neurometabolic coupling deficit from an inefficient mismatch between neural activity and hemodynamic recruitment. These competing hypotheses might be tested in future studies of SZ from anatomically precise measures of EEG parameters and neuroimaging markers of blood flow and/or metabolism. Hemodynamic response abnormalities identified in SZ (Ford et al., 2005) could reflect abnormal vasculature (Moises et al., 2015) or disrupted energy metabolism (Konradi and Öngür, 2017) leading to reduced BOLD correlation with gamma EEG and slope. SZ has been conceptualized as a deficit in bioenergetic coupling (Sullivan et al., 2018) leading to inefficient hemodynamic recruitment (Karch et al., 2009; Potkin et al., 2009; Schlösser et al., 2008; Zhou et al., 2016) that might yield a mismatch between EEG and BOLD signals.

### Implications and Future Directions

In single modality fMRI-studies, ongoing resting-state neural signals are mostly attributed to “internal” processes and the DMN (Raichle, 2015). Our focus on concurrent EEG-fMRI recordings reveals that EEG activity may also be linked to stimulus specific processing of the “external” environment. This processing might further interact with “internal” processing, or suggest individual differences in background processing, as we have observed within this population of SZ participants. Future studies might query participants about their experience in the scanner to derive post-hoc measures of attentional focus (e.g. scanner sounds or self-referential). We hope that our work prompts a re- consideration of resting-state methodology in SZ. Future studies might examine if well studied DMN connectivity differences might be impacted by participant attentional orientation while at rest (Benjamin et al., 2010).

### Limitations

Our study is primarily limited by the fact that we do not have unique and specific recordings of scanner sound onset/offset timing for direct correlation with EEG or BOLD signals on a participant by participant basis. Future researchers could potentially use scanner-background sound as a “stimulus” to examine auditory processing and sensory gating in a semi-naturalistic environment. While our results strongly suggest that EEG-BOLD coupling plays a role in auditory processing, our paradigm was not specifically designed to test this hypothesis. Although we did not find group differences in resting-state aperiodic slope or gamma power, we cannot rule-out the possibility that such differences would be present during a single modality EEG recording conducted outside of the scanner. For this reason, it is difficult to interpret our results in the context of other resting-state studies, particularly resting-state EEG studies that are recorded outside of a scanner. Future studies are needed to investigate how the scanner environment, including scanner sound, posture and being in an enclosed space might influence EEG activity (Thibault et al., 2014). These factors might differentially impact participants with SZ, indicating the need for detailed study of environmental scanner effects across clinical populations.

## Data Availability

Data and code are available from the authors upon reasonable request. Our informed consent procedure specified that de-identified data may be shared with collaborators. Our institutional policy requires that collaborators enter into a formal data sharing agreement with the San Francisco Veterans Affairs Health Care System.

## Acknowledgements and Disclosures

This work was supported by grants from National Institute of Mental Health (R01MH058262- 17 to JMF) and the VA (I01CX000497-06 to JMF, IK1CX002089 to MSJ). DHM consults for Boehringer Ingelheim. The authors have declared that there are no conflicts of interest in relation to the subject of this study.

